# Retrospective Comparison of Outcomes and Cost of a Virtual vs. Center-Based Cardiac Rehabilitation Program

**DOI:** 10.1101/2024.05.05.24306717

**Authors:** Neil D. Shah, Conor W. Banta, Andrea L. Berger, Andrea Hattenberger, Alex Zimmerman, Bryan E. Martin, Edward Wu, Usnish Majumdar, H. Lester Kirchner, Martin E. Matsumura

## Abstract

**Background:** Despite robust evidence supporting an association with improved outcomes in eligible patients, cardiac rehabilitation (CR) remains underutilized, with a minority of eligible patients participating. Virtual cardiac rehabilitation (VCR) has been proposed as an alternative to traditional center-based CR (CBCR) to improve utilization rates. However, data supporting the efficacy and cost-effectiveness of VCR is limited. In the present study, we compared the outcomes and cost of a VCR vs. traditional CBCR program.

**Methods:** Retrospective cohort study comparing VCR vs. CBCR. CBCR data were collected from a period of January 2018 to September 2023. VCR data were collected from program initiation in July 2021 to September 2023. Primary health outcomes measured were 1-year mortality rates, recurrent MI, all-cause hospital readmission, and emergency department visits. Primary cost outcomes were analyzed as cost ratios related to VCR vs. CBCR assessing total medical costs allowed, pharmacy costs, and total costs of care over the 12 months post-CR enrollment.

**Results:** VCR was associated with a significant reduction in 1-year all-cause hospital readmission (incident rate ratio (IRR)=0.616, 95% confidence interval (CI): 0.489, 0.777, p<0.001) and ED admission (IRR=0.557, 95% CI: 0.452, 0.687, p<0.001) at 1 year. The IRR of MI and all-cause mortality did not significantly differ between VCR and CBCR. In addition, VCR was associated with significant reductions in medically related (cost ratio=0.814, 95% CI: 0.690, 0.960, p=0.0144) and total costs (cost ratio=0.838, 95% CI: 0.725, 0.970, p=0.0176).

**Conclusion:** VCR is a viable alternative to CBCR with at least comparable efficacy and cost, and as such, represents a key mechanism for improving access to and participation in CR for eligible patients.

## INTRODUCTION

Cardiac rehabilitation (CR) is a structured exercise and counseling program focused on the secondary prevention of cardiovascular disease (CVD). CR has been demonstrated to drive robust improvements in rates of all-cause mortality and rehospitalization via improvements in physical fitness, heart function, and mental health^1,2^. Accordingly, CR is a Class 1 recommendation by the American Heart Association and American College of Cardiology (AHA/ACC) for patients with acute coronary syndromes (ACS), patients who undergo coronary revascularization (including in non-ACS settings and after coronary bypass grafting), as well as a Class 2a recommendation for those with symptomatic heart failure.

Despite broad applicability, CR remains underutilized. A recent national study of Medicare enrollees found that only 28.6% of eligible patients started a CR program, and only 7.9% completed a full 36-session CR program^3^. The Million Hearts Initiative, a program of the Centers for Disease Control, targets 70% CR participation, a figure that cannot be achieved without significant changes to the healthcare delivery ecosystem^4^. While behavioral and educational interventions have been developed to improve the rate of referral^5,6^, there remain significant structural challenges to overcome - availability of CR facilities, distance to CR facility, out of-pocket costs, lack of transportation, and language barriers^7^. A recent modeling study estimated that if all center-based CR facilities operating before the COVID-19 pandemic operated at maximal capacity, only 47% of eligible US patients could be accommodated^8^. Furthermore, a recent geographic survey revealed that approximately 14% of US adults older than age 65 live in a CR facility “desert,” with untenable ratios of eligible patients to existing facilities^9^.

These access and capacity barriers were worsened by the COVID-19 pandemic, which led to the closures of approximately 8% of all US CR facilities^10^. Post-pandemic, there remains a sustained 14% decrease in CR utilization as compared to prior^11^. These declines in utilization were most profound among patients residing in rural areas and in vulnerable communities, which further compounded existing disparities in CR utilization along lines of social disadvantage^12-14^.

Several alternative CR delivery models have emerged as potential solutions to the problem of limited access to traditional center-based CR (CBCR). The majority of these models utilize digital technologies to augment or replace components of CBCR in the patient’s home. While virtual CR (VCR) programs benefit from flexibility in delivering traditional CR exercise sessions, this flexibility presents an opportunity to offer a more comprehensive approach to secondary CVD prevention, with many models including counseling on nutrition, medication adherence, comorbid mental health conditions, and tobacco cessation^15^. Previously described VCR programs at the Veterans Affairs Health System and Kaiser Permanente Southern California, for example, involve self-guided exercise sessions completed in the home, risk factor-oriented counseling delivered via a series of telephonic or audiovisual interactions with clinical staff, and basic biometric monitoring via mobile app or smartwatch^16,17^. Elsewhere, “hybrid” models have emerged, combining exercise sessions performed at a CR facility with sessions performed at home^18^. Virtual CR (VCR) involves direct clinical supervision of exercise sessions performed at home via synchronous audio-visual technology^15^.

In the US, the Veteran’s Affairs Healthy Heart Home-Based CR (HBCR) program has demonstrated reductions in all-cause mortality among those participating as compared to those not participating in the program^16^. The magnitude of this reduction may be comparable to prior clinical trials of CBCR. A cohort study of patients in Kaiser Permanente’s Southern California VCR program revealed a 21% lower readmission rate vs. those who underwent CBCR^17^. A similar cohort study at the University of California, San Francisco found that functional outcomes were comparable between VCR, CBCR, and a hybrid CR model^19^. While these non-CBCR models present opportunities to innovate and extend capacity, there remains limited data directly comparing the efficacy of HBCR to CBCR, especially when leveraging VCR.

In the present study, we evaluated a novel VCR program across the Geisinger Health System, a large regional health system serving both urban and rural patients throughout Pennsylvania. Geisinger’s CBCR program was temporarily closed during the COVID-19 pandemic, during which a VCR program was launched. Subsequently, Geisinger’s CBCR program was re-launched in 2022 and operated alongside the VCR program. In this pragmatic community context, we aim to compare the clinical efficacy and cost of VCR and CBCR.

## METHODS

### Study Design and Data Source

This retrospective cohort study across Geisinger Health System compared VCR and CBCR. Geisinger Health System serves a large portion of central and northeastern Pennsylvania with urban and rural settings, diverse socioeconomic backgrounds, and variable access to hospitals where CBCR programs are located. During the COVID-19 pandemic, CBCR programs were closed, and VCR became available in July 2021. The study investigated the impact of VCR on common cardiovascular disease risk factors and clinical outcomes as compared to CBCR. CBCR data were collected from a period of January 2018 to September 2023. VCR data were collected from its initiation in May 2021 to September 2023. CBCR data, including cohorts before and after VCR implementation, were pooled due to the similarity in baseline characteristics. Prospective data were collected on patients enrolled in Geisinger CR via electronic medical records, individualized treatment plans (ITP), and VCR session data.

The study was approved by the Geisinger Health System IRB. A waiver of informed consent was obtained due to the retrospective nature of the study design.

### Study Population

Patients were included in the study if they were ≥ 18 years of age and had a qualifying diagnosis for enrollment into CR, including acute myocardial infarction within the preceding 12 months, coronary artery bypass grafting, current stable angina pectoris, heart valve repair or replacement, percutaneous transluminal coronary angioplasty (PTCA) or coronary stenting, heart or heart-lung transplant, stable CHF with left ventricular ejection fraction ≤ 35%, or other cardiac conditions as specified through Medicare National Coverage Determinations. Exclusion criteria included severe arthritis, dementia, Parkinson’s, and hypertrophic cardiomyopathy. Patients were excluded if they did not complete at least one session or had interruptions in their CR series which resulted in CR sessions spanning beyond 1 year following the enrollment date. For patients who were enrolled more than once into CR during the study period, only the initial CR series was included in the analysis.

### Outpatient Cardiac Rehabilitation Programs

Patients were referred to cardiac rehabilitation through a provider-placed referral order or via chart review identifying patients with eligible diagnoses. From January 2018 to March 2020, patients referred for CR were offered CBCR. From March 2020 to June 2022, CBCR locations were closed due to the COVID-19 Pandemic, and VCR was offered as the sole CR modality from July 2021 to June 2022. Starting in June 2022 until the end of the data collection period in September 2023, both options were offered, and providers were permitted to specify a request for VCR, CBCR, or no preference. For the latter period, if a provider preference was not stated on the referral order, patients were presented with both options. All referrals were reviewed for CR exclusion criteria and adequate insurance coverage for their selected CR modality.

The Geisinger VCR program is a partnership with Recora (New York, NY). Recora’s VCR platform is a comprehensive virtual program that includes both technology and services. After establishing a VCR consult, patients were mailed a kit including an internet-connected tablet with the Recora app preloaded, blood pressure and heart rate monitor, and exercise bands. Patients also had the choice to use their own smart device and download the app via the Apple App Store or Google Play Store. Patients were then onboarded via a real-time video visit followed by up to 36 real-time video sessions, including monitored group exercise sessions, nutrition counseling, risk factor modification, and psychosocial/wellbeing discussions. Sessions occurred 2-3 times per week for 12-18 weeks. Non-physician staff included exercise physiologists, dieticians, and ancillary staff, all provided by Recora. A standardized ITP was developed at the first visit and updated every 30 days until program completion. A Geisinger cardiologist was “available and accessible for medical consultations and emergencies” as part of virtual supervision responsibilities. The program concluded with a final set of exercise tests and questionnaires to document progress.

The Geisinger CBCR program operates across multiple Geisinger sites. The program is structured to include three sessions per week for a total of 12 weeks using a traditional CBCR structure based on guidelines of the American Association of Cardiovascular and Pulmonary Rehabilitation. This program consists of one-hour sessions, three times per week. The personnel present during cardiac rehab sessions are an exercise physiologist and a registered nurse who are present for the duration of the session. There is a medical director who oversees the administration of the program. The program duration target is 36 sessions.

### Outcome Measures

Primary health outcomes measured beginning from the first CR session were: 1-year mortality, recurrent MI, all-cause hospital readmission, and emergency department visits. Primary cost outcomes were total medical costs allowed, pharmacy costs, and total costs of care over the 12 months post-CR enrollment.

### Statistical Analysis

Descriptive statistics including mean and standard deviation for continuous variables, and frequency percentages for categorical variables are reported. Demographic and clinical characteristics of the sample were presented and stratified by program type. Baseline characteristics between programs were compared using Pearson’s Chi-square and Wilcoxon Rank-sum tests. Since this is a non-randomized study, we expected some baseline variables to vary between programs. To account for the imbalance (i.e., confounding) we estimated the propensity score (PS) for being in VCR using logistic regression. The PS was then used in the analysis to weight observations to create a population balanced on all baseline variables. Balance was measured using standardized mean differences (SMD). SMD < 0.10 is considered balanced. See Table 1 for the list of covariates used in the PS model.

**Table 1:** Cohort Demographic and Clinical Characteristics Stratified by Program Type.

|  | CBCR<br>n=2303 | VCR<br>n=703 | p |
| --- | --- | --- | --- |
| Age (yrs), mean (SD) | 66.9 (11.35) | 70 (10.83) | <0.0001 |
| Female sex, n (%) | 749 (32.4) | 269 (38.3) | 0.0031 |
| Race, n (%) |  |  |  |
| White | 2261 (98.2) | 673 (96.8) | 0.0016 |
| Non-white | 41 (1.8) | 22 (3.2) |  |
| Ethnicity, n (%) |  |  |  |
| Non-Hispanic | 2278 (99.4) | 675 (97.5) | <0.0001 |
| Latino/Hispanic | 14 (0.6) | 17 (2.5) |  |
| Hx HTN, n (%) | 1427 (61.7) | 546 (77.7) | <0.0001 |
| Hx Hyperlipidemia, n (%) | 1311 (56.7) | 490 (69.7) | <0.0001 |
| Hx Diabetes, n (%) | 664 (28.7) | 268 (38.1) | <0.0001 |
| Hx Myocardial infarction, n (%) | 1044 (45.1) | 288 (41.0) | 0.1441 |
| Hx Coronary artery disease, n (%) | 1538 (66.5) | 602 (85.6) | <0.0001 |
| Hx Congestive heart failure, n (%) | 445 (19.2) | 204 (29.0) | <0.0001 |
| Hx Chronic kidney disease, n (%) | 751 (32.5) | 335 (47.7) | <0.0001 |
| Alcohol abuse, n (%) | 93 (4.0) | 40 (5.7) | 0.0295 |
| Anemia/nutritional disorder, n (%) | 392 (17.0) | 188 (26.7) | <0.0001 |
| Iron deficiency, n (%) | 60 (2.6) | 23 (3.3) | 0.5571 |
| Leukemia, n (%) | 18 (0.8) | 5 (0.7) | 0.9198 |
| Lymphoma, n (%) | 23 (1.0) | 15 (2.1) | 0.0667 |
| Metastatic cancer, n (%) | 18 (0.8) | 12 (1.7) | 0.1020 |
| Solid tumor in situ, n (%) | 56 (2.4) | 23 (3.3) | 0.5571 |
| Solid tumor, malignant, n (%) | 232 (10.1) | 109 (15.5) | <0.0001 |
| Cerebrovasc disease, n (%) | 373 (16.2) | 127 (18.1) | 0.4940 |
| Coagulopathy, n (%) | 217 (9.4) | 96 (13.7) | 0.0023 |
| Dementia, n (%) | 61 (2.6) | 32 (4.6) | 0.0336 |
| Depression, n (%) | 469 (20.4) | 164 (23.3) | 0.1668 |
| Diabetes w/ chronic comp, n (%) | 590 (25.6) | 251 (35.7) | <0.0001 |
| Diabetes w/o chronic comp, n (%) | 807 (35.0) | 302 (43.0) | <0.0001 |
| Drug abuse, n (%) | 41 (1.8) | 19 (2.7) | 0.0010 |
| HTN, uncomplicated, n (%) | 1756 (76.2) | 599 (85.2) | <0.0001 |
| HTN, complicated, n (%) | 807 (35.0) | 341 (48.5) | <0.0001 |
| Heart failure, n (%) | 693 (30.1) | 282 (40.1) | <0.0001 |
| Liver disease, mild, n (%) | 202 (8.8) | 96 (13.7) | <0.0001 |
| Liver disease, mod to sev, n (%) | 16 (0.7) | 7 (1.0) | 0.0623 |
| COPD, n (%) | 530 (23) | 224 (31.9) | <0.0001 |
| Neurologic/movement dis, n (%) | 77 (3.3) | 30 (4.3) | 0.1126 |
| Seizures/epilepsy, n (%) | 63 (2.7) | 23 (3.3) | 0.5565 |
| Paralysis, n (%) | 69 (3.0) | 19 (2.7) | 0.5819 |
| Obesity, n (%) | 972 (42.2) | 341 (48.5) | <0.0001 |
| Periph vasc disease, n (%) | 659 (28.6) | 297 (42.2) | <0.0001 |
| Pulm circulation disease, n (%) | 182 (7.9) | 59 (8.4) | 0.0505 |
| Renal failure, moderate, n (%) | 512 (22.2) | 233 (33.1) | <0.0001 |
| Renal failure, severe, n (%) | 79 (3.4) | 43 (6.1) | <0.0001 |
| Hypothyroidism, n (%) | 410 (17.8) | 157 (22.3) | 0.0013 |
| Other thyroid disorders, n (%) | 128 (5.6) | 59 (8.4) | <0.0001 |
| Peptic ulcer with bleeding, n (%) | 62 (2.7) | 24 (3.4) | 0.5549 |
| Valvular heart disease, n (%) | 657 (28.5) | 269 (38.3) | <0.0001 |
| Weight loss, n (%) | 132 (5.7) | 53 (7.5) | 0.0623 |

After balancing the baseline covariates, the health outcomes were assessed using a weighted time-to-event analysis to estimate the incident rate (#events/10,000 patient days of follow-up). Weighted Gamma regression models that included an offset for the number of follow-up months were used for the cost outcomes. Results are reported as annual incident rate and incident rate ratio with 95% CIs for the health outcomes and the ratio of mean per member per month costs expressed as VCR:CBCR and 95% CIs for costs. The comparison of costs between groups was accomplished by testing the ratio of the averages due to the type of CR program. Statistical analyses were performed using the twang package^20^ in R v4.3.3^21^ and SAS v9.4 (SAS Institute Inc., Cary, NC, USA). For all comparisons p<0.05 was considered significant.

## RESULTS

There were 3006 patients who participated in CR with mean [SD] age, 67.7 [11.3] years, 1017 [33.8%] women and 2924 [97.9%] were white. A total of 2303 patients were enrolled in CBCR (76.6%) and 703 in VCR (23.4%). In addition to being significantly older, the VCR group had significantly higher rates of hypertension, hyperlipidemia, diabetes, coronary disease, heart failure, valvular disease, peripheral vascular disease, kidney disease, and chronic pulmonary disease (Table 1). After weighting of the observations using the PS, all covariates were balanced (all SMD <0.10). Patients enrolled in VCR completed a significantly higher median number of CR sessions vs. CBCR (34.4 vs. 21.4, p<0.001, data not shown).

Propensity-weighted health outcomes of CBCR and VCR are presented as 1-year incidence rate and incident rate ratio (IRR). VCR was associated with a statistically significant reduction in 1-year all-cause hospital readmission (14.9 vs 24.2 events, IRR=0.616, 95% CI: 0.489, 0.777), p<0.001) and ED admission (26.8 vs. 48.0 events, IRR=0.557, 95% CI: 0.452, 0.687, p<0.001). There were no significant differences in myocardial infarction and overall mortality between CBCR and VCR consistent with prior studies^17,25^. (Table 2) It was also found that those in VCR completed an average of 34.4 CR encounters, whereas those in CBCR had an average of 21.4 CR encounters (p<0.001)

**Table 2:** Weighted Analysis of Health Outcomes at 1 Year Post-Enrollment.

|  |  | CBCR (n=2303) | VCR (n=703) | Incident Rate Ratio | P-Value |
| --- | --- | --- | --- | --- | --- |
| Inpatient readmission | Number of Events | 444 | 118 | 0.616<br>(0.489, 0.777) | <0.001 |
|  | Annual Incident Rate | 24.2 (21.9, 26.7) | 14.9 (12.1, 18.4) |  |  |
| ED admission | Number of Events | 806 | 150 | 0.557<br>(0.452, 0.687) | <0.001 |
|  | Annual Incident Rate | 48.0<br>(44.5, 51.8) | 26.8<br>(22.0, 32.5) |  |  |
| Myocardial infarction | Number of Events | 30 | 10 | 1.250<br>(0.535, 2.914) | 0.605 |
|  | Annual Incident Rate | 1.4<br>(0.1, 2.1) | 1.8<br>(0.8, 3.7) |  |  |
| Mortality | Number of Events | 57 | 16 | 0.618<br>(0.345, 1.104) | 0.104 |
|  | Annual Incident Rate | 3.3<br>(2.5, 4.3) | 2.0<br>(1.2, 3.4) |  |  |

The cost analysis was restricted to patients covered by the Geisinger Health Plan (GHP) to allow collection of claims data. There was a total of 220 VCR and 375 CBCR patients that had GHP insurance when they were enrolled in CR. Table 3 reports the weighted costs comparisons. Total medical allowed and total costs allowed were found to be significantly reduced in the VCR group. More specifically, total medical allowed in the VCR group was observed to have an 18.6% reduction in the average per member per month (PMPM) (95% CI: 0.690, 0.960, p=0.0144). Similarly, the total costs allowed resulted in 16.2% lower average PMPM (95% CI: 0.725, 0.970, p=0.0176)

**Table 3:** Weighted Analysis of Cost Outcomes at 1 Year Post-Enrollment.

|  | Cost Ratio (VCR/CBCR) | P-value |
| --- | --- | --- |
| Total Medical Costs | 0.814 (0.690, 0.960) | 0.014 |
| Total Prescription Costs | 0.928 (0.753, 1.143) | 0.480 |
| Total Cost of Care | 0.838 (0.725, 0.970) | 0.018 |

## DISCUSSION

In the present study, we retrospectively compared outcomes of a novel VCR program and a traditional CBCR program using a propensity matching strategy to control for bias introduced by a retrospective observational study design. Our results suggested that compared to CBCR, VCR was associated with a significantly lower rate of all-cause rehospitalization and emergency department utilization at 1 year following initiation of CR sessions. These improved outcomes were associated with lower overall total cost of care for VCR participants. Our study adds to the existing body of data supporting the concept that VCR is a cost-effective strategy to deliver CR with patient outcomes at least comparable to traditional CBCR. Prior work has examined the efficacy and cost-effectiveness of VCR vs. CBCR. Nkonde-Price et al demonstrated a significant reduction in 1 year all-cause hospitalization in a retrospective cohort study of VCR vs. CBCR in the Kaiser Permanente system^17^. Oehler et al noted a nonsignificant trend toward cost savings associated with VCR compared to CBCR; however, hospitalization and emergency department usage were not different between CR modalities^22^. To the best of our knowledge, our study is the first to demonstrate a statistically significant improvement in outcomes associated with VCR vs. CBCR coupled with a favorable trend in overall healthcare costs among patients participating in VCR.

Placing the present study in context with available studies assessing the efficacy of VCR vs. CBCR is challenging due to the heterogeneous nature of VCR programs. Meta-analyses comparing CBCR and remote CR programs are difficult to generalize as they combine a multitude of delivery models and include scant data from the United States, where different barriers are likely to exist with regard to access to digital technologies and participation in exercise sessions^23^. Existing data on the comparative efficacy of VCR, which delivers a core CR experience closest to CBCR, is limited to low- and moderate-risk patients, with little coverage of patients with multiple comorbidities, who are > 75 years of age, women, or who belong to minority racial and socioeconomic groups^24^. Differing from the existing data and a novel aspect of this study, was the higher medical complexity and number of comorbidities in the patients enrolled in VCR as compared to CBCR. As our study utilized a unique VCR program developed and delivered by a third-party entity, no assumption can be made that similar improvements in outcomes would be seen using alternative and less comprehensive programs. However, it is important to note that our study adds to the data that a VCR program utilizing smart devices and synchronous video visits is feasible in an older patient population that tends to have limited technology literacy. This was in agreement with Harzand et al who showed the feasibility of a smartphone approach to cardiac rehab in an older veteran population^25^.

In tandem with clinical efficacy, the cost-effectiveness of VCR models is another important factor in their feasibility. A recent meta-analysis of 9 different VCR programs found that 7 were likely cost-effective, though again these findings are difficult to generalize due to the heterogeneity in CR delivery models and country-level differences between programs^26^. A recent study found that asynchronous CR may be more cost-effective to deliver than traditional CBCR, when driving comparable health outcomes^22^. However, asynchronous CR may face limited uptake given its lack of supervision and clinician feedback during sessions. The present study adds to the available evidence that VCR can be associated with favorable overall patient cost while delivering an efficacious CR program.

The driver of improved outcomes of VCR in our study is unknown but several possibilities can be hypothesized. First, the intensity and comprehensive nature of a synchronous VCR program may play a role. The Recora program allows a focus on lifestyle modifications beyond the limited interactions during CR sessions. The application allows patients to log vitals, meals, weight, daily steps, physical activity, and mood. Tracking these parameters on a frequent basis rather than only at CBCR sessions could have led to improved outcomes through better patient awareness. Specifically, the availability of home blood pressure and wearable physical activity monitors may have been beneficial as shown in a meta-analysis by Hannan et al that concluded wearable physical activity monitors were superior to no device in improving cardiorespiratory fitness in the maintenance phase of CR^27^.

A second possibility is that the benefit may be related to the dose of CR afforded by VCR vs. CBCR. Patients utilizing VCR in our study completed 60% more sessions than those participating in CBCR. Prior studies support the concept that CR, like many interventions in cardiology, demonstrates a “dose-response” relationship, lending evidence to support this theory^28,29^. As the Geisinger Health System serves patients in a very large catchment area, it is possible that the dose of CR afforded by VCR was much higher than CBCR due to the ease of access and reduced barriers to attendance. Finally, the possibility of an undefined confounder related to the non-randomized nature of the study cannot be excluded.

Virtual and telehealth services have proliferated in recent years, and these modalities have improved access across a wide spectrum of clinical services and specialties. However, for continued broad adoption, telehealth will need to demonstrate both cost-effectiveness and clinical efficacy. Of note, the effects of telehealth on quality and Medicare spending have been recently studied, revealing a slight increase in spending and small improvements in quality^30^. The present study supports VCR as a clinical service that may be particularly well suited to telehealth delivery, given gains in access, clinical outcomes, and cost of care reductions.

The study has several limitations. The ideal method of comparison of outcomes of VCR vs. CBCR would be a randomized trial design. The present study was a retrospective, non-randomized observational study, and as such bias cannot be completely eliminated. It is possible that the discretion left to the ordering provider to select a mode of rehabilitation favored healthier patients enrolling in VCR rather than CBCR. Our robust efforts to risk match groups likely minimized but cannot eliminate this issue. It is notable that the outcomes of CBCR were very similar when comparing the study period prior to the introduction of VCR and during the period when patients were offered both VCR and CBCR. This consistency adds some degree of strength to the retrospective cohort design and results regarding the favorable outcomes of VCR vs. CBCR. Additionally, this study was conducted at a large integrated healthcare system that may not make this study generalizable to other patient populations. However, the benefit of VCR may actually be blunted in this study given the several CBCR options available.

In conclusion, the present study found that VCR was associated with reduced rates of hospitalization and ED utilization with reduced overall costs as compared to traditional CBCR. These findings lend further support for VCR as an alternative to CBCR especially in areas of limited access to traditional center-based programs.

## Data Availability

Data will be supplied upon reasonable request to the authors

